# Real-world before-and-after evaluation of AI support for lung cancer diagnosis at three US lung nodule clinics

**DOI:** 10.1101/2025.11.20.25340677

**Authors:** K Adam Lee, Amit Borah, Garrett Fiscus, Lisa Petricca, Sarah Dean, Annika Rings, Catarina Santos, Lyndsey C Pickup, Omar Ibrahim

## Abstract

**Background:** Health systems and payers require evidence that artificial intelligence (AI)-enabled decision support improves care delivery. Integrating AI into lung nodule management pathways may streamline workflows, improve identification and triage of patients with pulmonary nodules, and enable earlier lung cancer diagnosis. Yet real-world evidence of clinical utility remains limited. This study evaluated the impact of implementing a commercially available AI software to support the lung nodule and lung cancer care pathway across three US health systems.

**Materials and Methods:** A real-world before-and-after study compared clinic activity during matched 12.5-month pre-implementation and post-implementation periods at three US lung nodule clinics. Primary outcomes were the number of new patients and the number of invasive procedures performed. Secondary outcomes were numbers of lung cancers and early-stage (stages 0–II) lung cancers, and time from baseline CT to diagnosis. Analyses used Wilcoxon signed-rank and Mann-Whitney U tests, with medians and interquartile ranges (IQRs) reported for continuous measures.

**Results:** Across sites, new patients increased from 138 before to 232 after implementation, equivalent to 11.0 to 18.6 patients per month (*p*=0.0002), with the median rising from 9 (IQR 7–14) to 16 (IQR 13–21) patients. Monthly invasive procedures increased from 5.0 to 7.8 (*p*=0.0093). Monthly cancers increased from 3.0 to 3.8 (*p*=0.3242), while monthly early-stage cancers rose from 1.4 to 2.4 (*p*=0.0996). Median time to diagnosis was 58 days (IQR 33–153) before and 57 days (IQR 27–85) after implementation (*p*=0.28), with IQR narrowing from 120 days to 58 days.

**Conclusion:** Implementation of the AI-enabled software was associated with significantly increased patient throughput resulting in an increase in lung cancer diagnoses. These results, observed prospectively post-implementation, suggest AI-assisted identification and triage can expand access to timely specialist review while maintaining diagnostic efficiency.

## Introduction

Lung cancer is the leading cause of cancer mortality worldwide [1], with most patients diagnosed at an advanced stage when curative treatment is no longer possible [2,3]. Early detection and management of pulmonary nodules are therefore critical to improving outcomes. Pulmonary nodules are frequently identified on chest computed tomography (CT) scans performed for a variety of clinical reasons [4,5]. Although most of these incidentally detected pulmonary nodules are benign, some represent early-stage lung cancer. International guidelines recommend management strategies based on estimated malignancy risk, using physician judgement alongside risk calculators such as the Mayo and Brock (PanCan) model [6–8]. However, implementation of these guidelines in routine practice across the United States (US) is inconsistent, with 50–60% of patients with incidental pulmonary nodules not receiving guideline-adherent follow-up, even before risk stratification, due to manual workflows and management systems [9–12]. Contributing factors include insufficient documentation in radiology reports; communication breakdowns between care providers; the lack of centralized, automated tracking systems; and unclear accountability for managing incidental findings [13]. These challenges affect the lung nodule care pathway, from case identification through to risk assessment, follow-up, and diagnostic evaluation.

Patients with indeterminate or intermediate-risk nodules pose the greatest clinical challenge, as management may involve either continued surveillance or invasive diagnostic procedures [6]. As such, it is important that such nodules are followed up and managed in a clinically appropriate and timely manner.

Artificial intelligence (AI) software tools have been developed to address these challenges and support key steps in the lung nodule care pathway. Some of these tools focus on CT image analysis, providing image-based detection (computer-aided detection; CADe), classification, risk stratification, and malignancy prediction (computer-aided diagnosis; CADx) to standardize assessment of indeterminate pulmonary nodules and support guideline-adherent clinical decision-making [14–19]. Others apply natural language processing (NLP) to radiology reports to automatically and systematically identify and track patients, helping to improve capture of patients with incidental nodules, increase rates of appropriate follow-up, and reduce missed or delayed diagnoses [20–23].

Despite this emerging evidence, there is limited multi-site, real-world evaluation of the clinical utility of these AI software tools. Most published studies are limited to single centers or short evaluation periods, and assess measures of diagnostic performance and clinical management decisions, rather than pathway-level outcomes [14,16]. It remains unclear whether implementation of such platforms translates into measurable changes in patient volumes, diagnostic procedures, and time to diagnosis across diverse healthcare settings.

To help address this, we conducted a multi-site ambispective before-and-after evaluation of AI-enabled lung nodule and lung cancer pathways supported by one such FDA-cleared, commercially available AI software system. The software integrates automated identification, management, and tracking of pulmonary nodule patients from radiology reports, and combines this with an AI lung cancer prediction algorithm for CADx that analyzes CT images to support clinical decisions. We compared clinic-level activity across three US health systems before and after implementation of the AI software, measuring monthly patient volumes, procedures, cancer diagnoses (including stage at diagnosis), and time from baseline CT scan to diagnosis.

We aimed to generate real-world clinical utility evidence for health systems and payers on the impact of AI-enabled lung nodule and lung cancer pathways. To meet this aim, we assessed whether the integration of an AI software system is associated with improvements in the number of pulmonary nodule patients followed up and in the overall timeliness, throughput, and reliability of nodule care in routine practice. Scaled adoption has the potential to increase early lung cancer diagnoses through earlier and more reliable follow-up, while supporting efficient planning and evaluation of nodule services. The study achieved this aim, and the results offer practical evidence for clinicians, health system leaders, and payers.

## Materials and Methods

### Study design and setting

This was a multicenter ambispective real-world before-and-after observational cohort study conducted at three US health systems. The study was reported in accordance with the Strengthening the Reporting of Observational Studies in Epidemiology (STROBE) guidelines [24]. The participating sites were diverse: a regional health system hospital (site A), a community hospital (site B), and an academic medical center (site C). Each site implemented the AI software at a defined “go-live” date. The pre-implementation enrollment period at each site served as the baseline comparator, and the post-implementation enrollment period comprised the equivalent calendar months in the following year (Table 1). No additional staffing was required for the post-implementation period. Collecting data for paired months in this way helped mitigate seasonal variation. The specific pre-implementation and post-implementation enrollment periods are shown in Table 1, while patient numbers at each site, before and after exclusion criteria were applied, are shown in the study flow diagram (Fig 1).

**Table 1.**
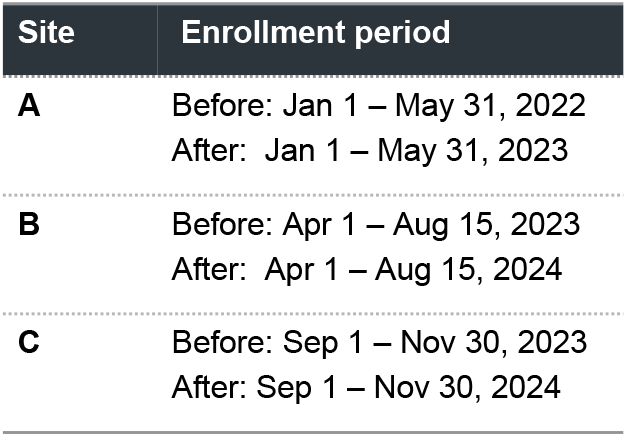
Enrollment periods before and after implementation.

| Site | Enrollment period |
| --- | --- |
| A | Before: Jan 1 – May 31, 2022<br>After: Jan 1 – May 31, 2023 |
| B | Before: Apr 1 – Aug 15, 2023<br>After: Apr 1 – Aug 15, 2024 |
| C | Before: Sep 1 – Nov 30, 2023<br>After: Sep 1 – Nov 30, 2024 |

**Fig 1.**
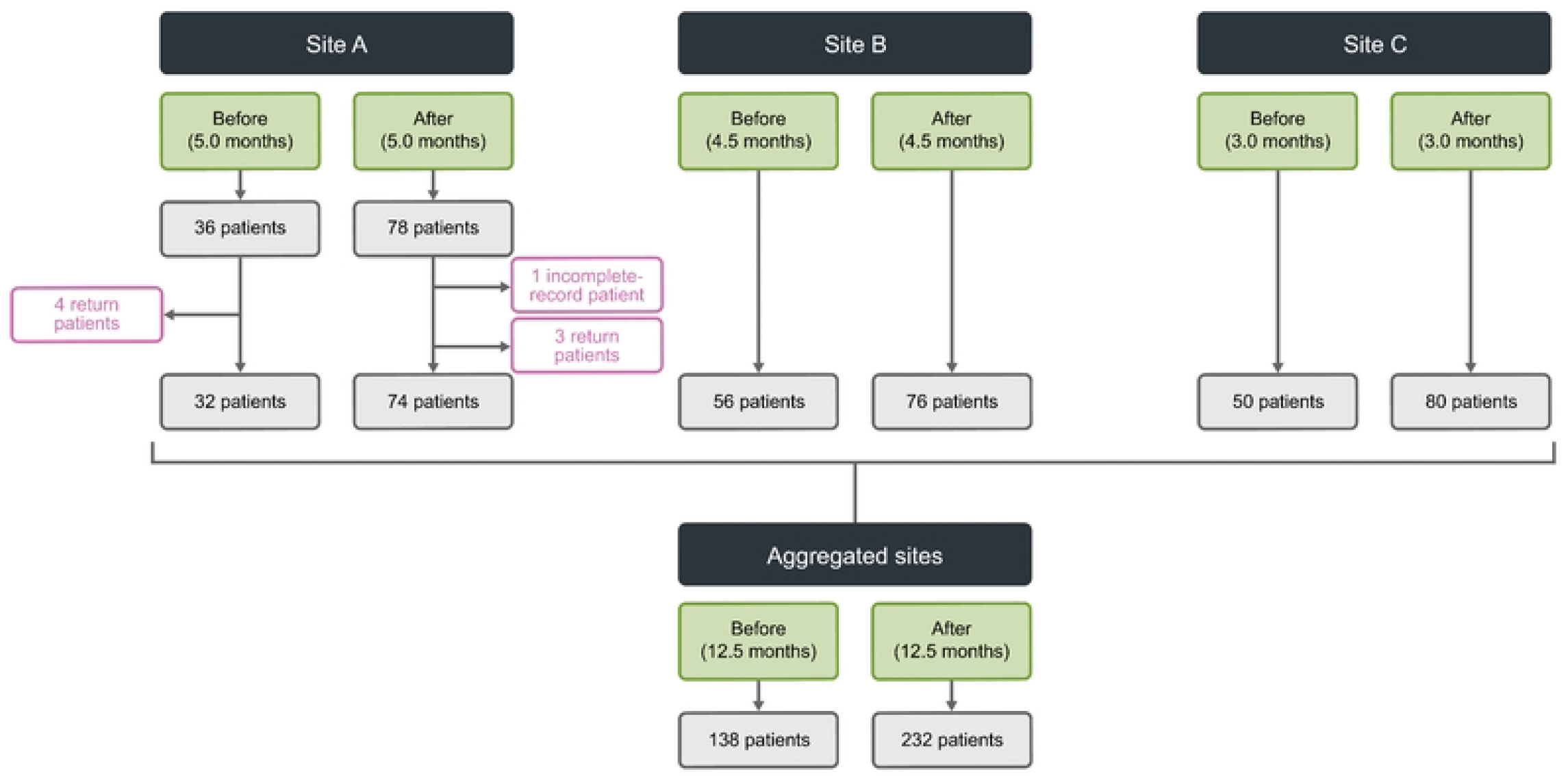
Study flow diagram across sites and periods.

### Study population

The study population included all consecutive new patients who attended the dedicated lung nodule clinics at the three sites during the predefined enrollment periods (Table 1). Eligible patients were those with incidentally detected pulmonary nodules on CT who had their first nodule-related clinic visit during the pre-implementation or post-implementation enrollment periods. Patients were excluded if they were return patients already under active follow-up before the study period, had incomplete data, or were enrolled outside of the defined enrollment periods.

### Intervention and comparator

The AI software deployed at the three sites and evaluated in this study was the Virtual Nodule Clinic (VNC), developed by Optellum Ltd (Oxford, United Kingdom). Site A used VNC version 2.3.2.3, site B used version 2.3.3.8, and site C used version 2.3.3.7. VNC is an FDA-cleared, web-based clinical decision support platform for pulmonary nodule management, classified as a Class II medical device. It integrates three modules: a patient discovery tool that uses NLP AI to automatically identify patients with pulmonary nodules from radiology reports; a CADx AI algorithm, Lung Cancer Prediction (LCP), that analyzes CT images to generate a malignancy risk score (from 1 to 10) to support consistent clinical decision-making; and a patient tracking and management dashboard that supports longitudinal follow-up and flags missed appointments. The LCP algorithm provides these quantitative, standardized risk estimates to augment physician judgement and guideline-based management, with previous retrospective studies validating its diagnostic performance and potential clinical utility in indeterminate pulmonary nodules [17,25–27].

During the post-implementation period, all sites implemented an AI-enabled pathway used to identify, risk-stratify, and track patients. The AI software automatically identifies patients with incidentally reported pulmonary nodules and adds them to a centralized dashboard. Clinicians then review the listed patients and, supported by LCP AI risk stratification, determine the appropriate clinical decision, whether that be discharge, surveillance, or intervention. Their decisions are recorded within the software to support the patient’s longitudinal care.

The comparator was the standard lung nodule and lung cancer care pathway prior to implementation, which relied on manual identification of incidental nodules and conventional tracking processes without VNC. Management decisions during both periods followed established guidelines and physician judgement [6,28].

### Data collection and outcomes

Data were extracted from electronic health records and nodule clinic databases at each site. For the pre-implementation period, data were collected retrospectively, whereas for the post-implementation period, data were collected prospectively as part of the VNC-supported workflow. Each patient’s record was reviewed for the date of baseline CT, clinic visits, diagnostic procedures, and outcomes. Outcomes were observed within a predefined follow-up window after the first clinic visit. For each site, the duration of this window was set by the post-implementation data-entry end date and that duration was applied equally to the pre-implementation cohort. The post-implementation data-entry end dates were: site A - May 31, 2023; site B - Dec 31, 2024; and site C – Aug 31, 2025.

The primary outcomes were the monthly volume of new patients attending the lung nodule clinic and the number of procedures performed, defined as percutaneous or bronchoscopic biopsies and surgical resections. Secondary outcomes included the number of confirmed lung cancers diagnosed, the number of early-stage lung cancers (stages 0–II), and the time from baseline CT to confirmed cancer diagnosis, measured in days.

Lung cancer diagnoses were confirmed by histopathology. In contrast, nodules were classified as benign if confirmed by histopathology, stable on imaging for at least 12 months, or resolved on follow-up. Cancer stage at diagnosis was extracted from pathology or oncology records. Time to diagnosis was defined as the number of days from the baseline CT that identified the nodule to the date of confirmed cancer diagnosis. For pre-implementation cases where the diagnosis occurred beyond the length of the matched post-implementation follow-up period, the case was censored from cancer- and procedure-related analyses to ensure an equal follow-up window for both pre- and post-implementation periods. For analyses of diagnostic timelines, patients whose time from baseline CT to confirmed lung cancer diagnosis exceeded two years (>730 days) were excluded, as such long delays were considered outliers not reflective of the clinical diagnostic pathway.

### Statistical analysis

Outcomes were summarized using descriptive statistics. Counts and proportions were reported for categorical variables, and medians with interquartile ranges (IQRs) for continuous outcomes. Monthly outcomes were analyzed as paired data, with each “after” month compared to the same calendar month one year earlier. The Wilcoxon signed-rank test was used to compare paired monthly outcomes (patients, cancers, early-stage cancers, procedures). For these site-level paired-month outcomes, period (before versus after implementation) was treated as a within-site repeated factor, with months paired one year apart. The Wilcoxon signed-rank test assumes paired observations with differences that are approximately symmetric; our within-site month pairs (same calendar month one year apart) satisfied these conditions. The Mann-Whitney U test was used to compare distributions of time to diagnosis between pre- and post-implementation cohorts. For this patient-level time-to-diagnosis analysis, period was treated as a between-subjects factor. The Mann-Whitney U test assumes independent groups and an ordinal or continuous outcome with similarly shaped distributions; these conditions were met. No multivariable models were fitted, so linearity and multicollinearity did not apply. All tests were two-sided, with statistical significance defined as p<0.05. No formal sample size calculation was performed, since all eligible patients within the predefined enrollment periods were included. Although data were collected beyond the paired months, site-specific analyses demonstrated seasonal variation; therefore, paired-month comparisons were adopted for the primary multi-site analysis, with additional sensitivity analyses by site and with outliers included to test the robustness of the results. Statistical analyses were performed using Python version 3.11.13 (packages: pandas 2.3.3, scipy 1.16.1, statsmodels 0.14.5).

### Ethics

Data were collected from routine clinical care. Institutional review boards at each participating site approved the study with a written waiver of informed consent and HIPAA authorization. All analyses were performed on de-identified data. Clinical authors at each site had access to identifiable patient information solely within their own institutions during data collection. All data were de-identified before being shared with authors external to the participating sites for analysis.

## Results

### Primary outcomes

#### Study population

Across three sites and 12.5 paired months per phase (5.0 at site A, 4.5 at site B, 3.0 at site C), 378 unique new patients attended lung nodule clinics in the enrollment periods: 142 at before implementation and 236 after implementation. After excluding seven return patients and one patient who had incomplete data, there were 138 patients before and 232 patients after implementation included in the final analysis (Fig 1). Details regarding numbers per site and the enrollment periods are provided in Fig 1 and Table 1, respectively.

#### Clinic volumes

The total number of new patients increased from 138 to 232 across paired months, equivalent to 11 to 18.6 patients per month (a 68% increase). The median monthly number of patients increased from 9 (IQR 7–14) before implementation to 16 (IQR 13–21) after implementation (Wilcoxon signed-rank statistic = 0.0, *p* = 0.0002), indicating a statistically significant 78% increase (Table 2). By site, monthly new-patient volumes increased from 6.4 to 14.8 (131% increase) at site A, 12.2 to 16.9 (38% increase) at site B, and 16.3 to 27.3 (68% increase) at site C.

**Table 2.** Primary and key secondary outcomes before versus after VNC implementation (paired months across three sites). Procedures correspond to percutaneous or bronchoscopic biopsies, or surgical resections. Early-stage cancer refers to cancers diagnosed at stages 0–II. CT, Computed tomography; IQR, Interquartile range; VNC, Virtual Nodule Clinic.

| Outcome | Before VNC implementation | After VNC implementation | Change | Statistical test and $p$-value from paired monthly data |
| --- | --- | --- | --- | --- |
| Median monthly new patients | 9 (IQR 7–14) | 16 (IQR 13–21) | +78% | Wilcoxon signed-rank ($p = 0.0002$) |
| Median monthly procedure volume | 5.0 | 7.8 | +57% | Wilcoxon signed-rank ($p = 0.0093$) |
| Monthly cancer diagnoses | 3.0 | 3.8 | +28% | Wilcoxon signed-rank ($p = 0.3242$) |
| Monthly early-stage cancer diagnoses | 1.4 | 2.4 | +56% | Wilcoxon signed-rank ($p = 0.0996$) |
| Median time from baseline CT to diagnosis | 58 (IQR 33–153) | 57 (IQR 27–85) | - | Mann–Whitney U ($p = 0.28$) |

#### Procedures

After censoring procedures that occurred outside the matched follow-up window, the number of patients undergoing at least one invasive diagnostic procedure (biopsy or surgical resection) increased from 62 to 97, equivalent to 5.0 to 7.8 procedures per month (57% increase; Wilcoxon signed-rank statistic = 9.5, *p* = 0.0093), indicating a statistically significant increase (Table 2). By site, monthly procedures rose from 2.2 to 4.8 (118% increase) at site A, from 5.6 to 7.1 (27% increase) at site B, and from 8.7 to 13.7 at site C (58% increase).

### Secondary outcomes

#### Cancer diagnoses

After censoring diagnoses outside the matched follow-up window, total confirmed lung cancer diagnoses increased from 37 to 48, equivalent to 3.0 to 3.8 cancers per month (28% increase; Wilcoxon signed-rank statistic = 21.5, *p* = 0.3242) (Table 2). Site-level monthly cancer counts increased from 0.6 to 2.0 (233% increase) at site A, decreased from 4.2 to 3.8 (10% decrease) at site B, and increased from 5.0 to 7.0 (40% increase) at site C.

#### Early-stage cancers

Restricting to lung cancer types of interest (non-small cell lung cancer [NSCLC], small cell lung cancer [SCLC], and carcinoid) and to cases with stage information available, early-stage cancer diagnoses (stage 0–II or limited) increased from 18 to 30 overall, equivalent to 1.4 to 2.4 per month (71% increase; Wilcoxon signed-rank statistic = 11.5, *p* = 0.0996) (Table 2). Stage at diagnosis was unknown for four of 28 pre-implementation lung cancers and four of 38 post-implementation lung cancers. Site-level early-stage counts with known stages increased from 0 to 8 at site A, increased from 10 to 11 (10% increase) at site B, and 8 to 11 at site C (37% increase).

#### Time to diagnosis

For time to diagnosis, cases that exceeded the two-year baseline-CT-to-diagnosis cutoff or occurred outside the matched follow-up window were censored. Among the cancer cases included in this time-to-diagnosis analysis, the median time from baseline CT to histologic diagnosis was 58 days (IQR 33–153) before and 57 days (IQR 27–85) after implementation (Mann-Whitney U = 949.5, *p* = 0.28) (Table 2). The difference was not statistically significant, although the IQR narrowed from 120 to 58 days, indicating more consistent diagnostic timelines after implementation. Site-specific medians and IQRs are shown in the boxplots in Fig 2.

**Fig 2.**
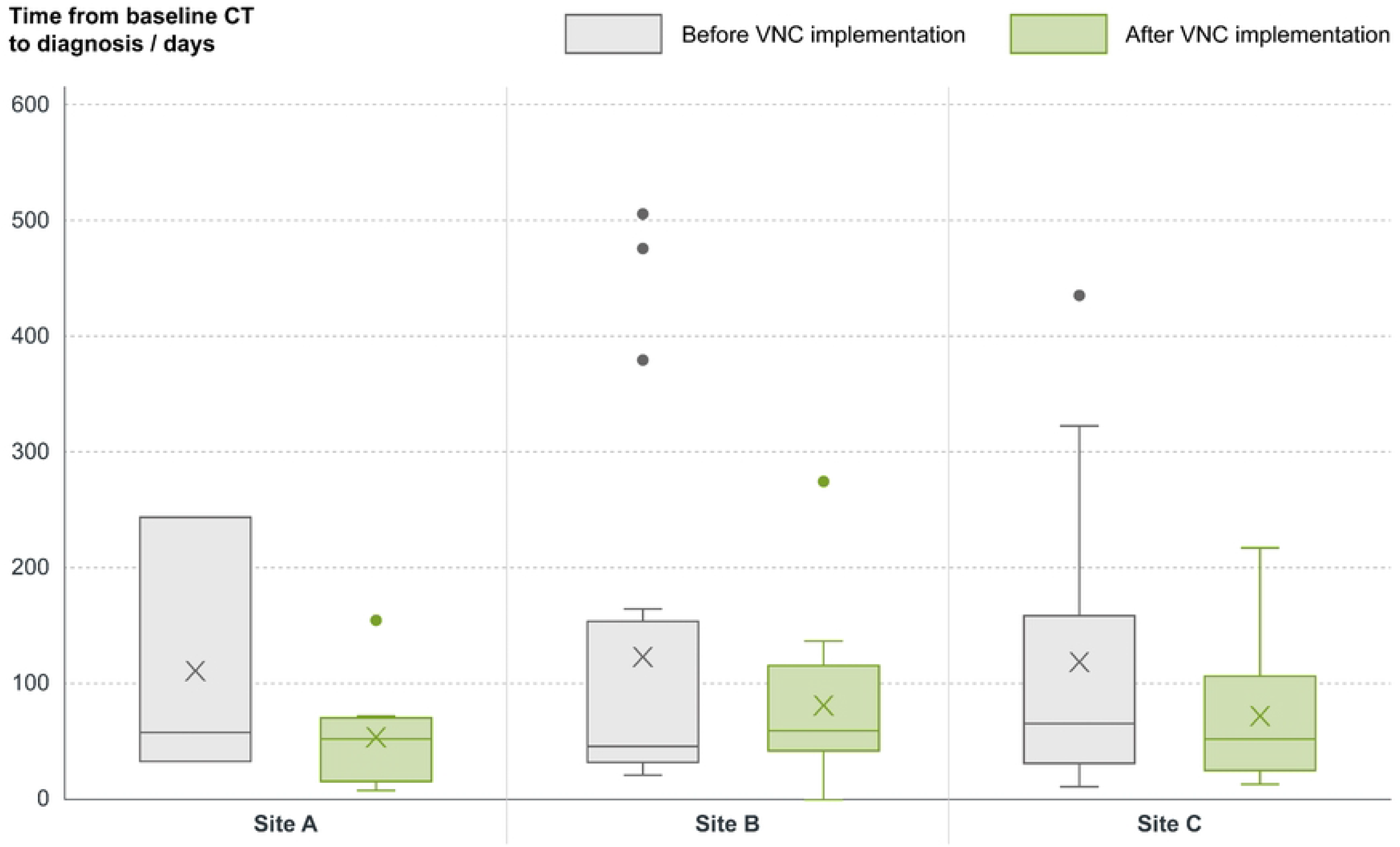
Time from baseline CT to confirmed lung cancer diagnosis before and after VNC implementation (paired months for each site). Boxplots show distributions of time to diagnosis in days, with each box representing data from paired-month analyses comparing the same calendar months before and after implementation. CT, Computed tomography; VNC, Virtual Nodule Clinic.

### Sensitivity checks

Including the outlier cases with a time from baseline CT to diagnosis greater than two years yielded medians of 59 days (IQR 35–160) before and 60 days (IQR 32–109) after implementation, with the same qualitative conclusion of no statistically significant difference and a narrower spread (from 125 to 77 days) after implementation. Results by site were directionally consistent with the primary analysis for clinic volumes, procedures, and time to diagnosis, with some heterogeneity expected across settings.

## Discussion

In this multi-site real-world ambispective evaluation, implementation of VNC and an AI-enabled lung nodule and lung cancer pathway was associated with significant changes in lung nodule clinic metrics. Across 12.5 paired calendar months, monthly new patients increased from 11 to 18.6 per month, a 68% increase, with the paired-month median increasing from 9 to 16 patients (Wilcoxon *p* = 0.0002). The volume of procedures increased from 5.0 to 7.8 per month, and this paired change was significant (*p* = 0.0093). These increases in the study’s primary outcomes were observed for each site. The most pronounced impact of the AI software was at site A, a clinic operated by the interventional pulmonology team at a regional health system hospital that had no prior system to capture lung nodule patients. At site A, new patient and procedure volumes rose 131% and 118%, respectively. Site B, a clinic run by a thoracic surgery team at a community hospital that previously relied on a simple email-referral system from the emergency department, also demonstrated meaningful but more moderate increases in new patient and procedure volumes (38% and 27%, respectively). Notably, these increases at site B occurred despite major pathway disruptions in the post-implementation period, including the loss of a key cancer referral source and the departure of the nurse navigator. These findings suggest that while all sites benefit from implementation, the magnitude of impact may differ according to real-world factors such as clinic capacity, available resources, existing workflows, and clinical specialty.

Cancer diagnoses rose from 3.0 to 3.8 per month, and early-stage cancers rose from 1.4 to 2.4 per month following implementation. Although these paired differences did not reach statistical significance (likely due to the limited sample size), the observed trend suggests potential clinical impact. Median time from baseline CT to confirmed cancer diagnosis was similar before and after implementation at 58 versus 57 days (Mann-Whitney U *p* = 0.28), while the IQR narrowed from 120 to 58 days, indicating more consistent timely follow-up. Taken together, clinics processed more patients without prolonging the time to diagnosis, with a signal toward more consistent timelines and more early-stage diagnoses.

These results extend prior evidence regarding the image-based risk stratification of the LCP algorithm. Multiple validation and reader studies have shown that LCP improves discrimination between malignant and benign nodules compared with clinical models, increases accuracy of clinician risk estimates, and reduces interobserver variability in management decisions [17,25–27]. In one such reader study of twelve experienced radiologists and pulmonologists, use of LCP increased diagnostic accuracy for all readers and by about 9% on average (AUC 0.82 to 0.89; *p* < 0.001); improved overall interobserver agreement for malignancy risk categories by 66% (κ = 0.35 versus κ = 0.58; *p* <0.001); and improved agreement on management recommendations by 18% (κ = 0.44 versus κ = 0.52; *p* = 0.001) [26]. These reader-level gains align with the system-level benefits we observed: by combining automated patient identification and tracking from radiology reports with LCP in a single workflow, VNC can operationalize steps that guidelines already recommend but that can be difficult to deliver reliably at scale [22]. For health systems and payers, the combination of higher throughput and earlier-stage diagnosis is noteworthy. Earlier stage treatment is generally less resource-intensive and associated with better outcomes [4,29], while avoidance of unnecessary follow-up and procedures in low-risk cases, if achieved, would reduce costs and patient burden [30].

The study has several strengths. First, it evaluates an FDA-cleared, commercially available AI software system implemented as part of the care pathway in three distinct practice environments, which supports external relevance. Second, outcomes reflect pathway performance and clinical utility, rather than diagnostic model performance alone. Monthly patient counts, procedures, cancer counts and stages, and times to diagnosis are directly interpretable by clinical users and payers. Third, the analysis strategy was prespecified, used paired calendar months to mitigate seasonality, excluded a ramp-up period, and applied non-parametric tests appropriate for operational data. Finally, the data gathered allow site-level inspection, with results that show comparable trends across academic, regional, and community clinics. These environments are representative of a broad spectrum of healthcare delivery in the United States, thereby enhancing the validity and generalizability of the results within organized nodule programs.

The study’s limitations, however, warrant cautious interpretation. First, the ambispective before-and-after design and absence of a contemporaneous control mean causality cannot be inferred, and residual confounding from changes in workflow, service availability, or staffing may have contributed to the observed effects. Second, the specific calendar months differed by site, which may introduce site-level temporal effects despite the paired-month approach. Finally, enrollment was limited to the paired months, and follow-up was assessed within site-specific matched follow-up windows. Longer observation may be needed to assess the significance and durability of any stage shift and the health-economic impact.

Several implications follow for practice and policy. Guideline-adherent diagnosis and management emphasize consistent, timely care for incidentally detected nodules [7,28]. Variation in adherence to follow-up recommendations remains common in real-world settings, and many patients receive delayed or no follow-up even before formal risk stratification [9].

Therefore, these findings support three desirable outcomes for care pathways and payers. First, earlier escalation for high-risk nodules, which can increase the volume of timely diagnostic procedures and shift diagnosis toward stages I and II, and, therefore, reduce downstream treatment costs and complications. Second, greater confidence in surveillance or discharge for low-risk nodules, which can reduce low-value follow-up and invasive testing. Third, the ability to manage a higher number of patients per month with similar or fewer staffing resources and without extending time to diagnosis, suggests improved operational efficiency, increased delivery of guideline-compliant follow-up, and reduced exposure to medico-legal risk. The earlier stage signal and tighter diagnostic timelines we observed align with those expectations. The rise in procedures likely reflects early adoption that favors sensitivity. As local teams gain experience and establish clear thresholds and governance for acting on LCP scores, specificity gains may be realized without sacrificing early lung cancer detection. Future research around the use of LCP and the wider VNC platform should investigate the health-economic impact of such AI software, and also focus on linking the LCP score more closely to guidelines, understanding how best to integrate it into clinical workflows to enhance decision-making and streamline care pathways.

## Conclusion

Deployment of VNC as part of an AI-enabled pathway in lung nodule clinics was associated with significant increases in patient volume and procedural activity. It was also associated with more consistent diagnostic timelines, resulting in an increase in lung cancer diagnoses. Taken together, these findings, observed prospectively across three diverse US health systems, suggest that AI-assisted case identification and triage can enhance access to timely specialist evaluation without compromising diagnostic efficiency. This supports the clinical utility of integrating AI-enabled clinical decision support software into lung nodule and lung cancer care pathways at scale.

## Data Availability

The study involves human research participant data containing potentially sensitive patient information. Legal and ethical restrictions, imposed by the participating institutions and by their Institutional Review Boards, prohibit public sharing of this data. Requests for data can be made by contacting the corresponding author.

## Acknowledgements

The authors thank J&J Interventional Oncology and Intuitive Surgical for funding the data collection, and the research teams at the sites for their diligent participation.

